# Meta-Research: Assessing the Alignment of Biomedical Research and Clinical Translation

**DOI:** 10.1101/2024.10.07.24314705

**Authors:** Mengyi Sun, Thomas Stoeger, Luís A Nunes Amaral

**Author notes:** Correspondence to: T.S., L.A.N.A.

## Abstract

More than half of later stage drug clinical trials fail due to lack of efficacy, highlighting gaps in our understanding of disease-target relationships. While it is widely believed that scientific research is essential for identifying drug-targets, whether scientists can allocate their research attention according to the clinical potential of disease-target relationships is remains a matter of debate. To address this knowledge gap, we analyzed data from Open Targets, a comprehensive disease-target relationship platform. We build an automated open pipeline that follows drugs through consecutive clinical trial phases and use it to uncover that research attention in the disease-specific primary literature has some predictive power for trial success. However, research attention toward specific targets are not predictive for overall trial success across diseases. Our study thus highlights the crucial, yet complex, role of the primary research literature in steering clinical trials, encouraging a more nuanced approach to translational medicine.

## Introduction

Drug development stands as the frontier of human progress, harnessing the power of scientific discovery to combat disease, extend lifespans, and improve quality of life. Central to this process is the identification of target genes. This is based on the principle that a drug can change a disease’s course by interacting with the molecules encoded by specific target genes involved in the disease’s onset or progression (1). *Infliximab* is a canonical example of the power of this principle (2): an antibody designed to target TNF-alpha, whose overproduction results in severe autoimmune responses. *Infliximab* thus ameliorates a wide range of autoimmune diseases, such as Crohn’s disease, rheumatoid arthritis, and psoriasis (2). Sadly, the development of *Inflixmab* is the exception rather than the norm — more than half of stage 3 and 4 drug clinical trials fail due to lack of efficacy (3) and only one out of ten drugs now entering phase 1 clinical trials will ultimately reach market approval (4).

The low success rate of new drug development pipelines is a topic of much discussion, and many potential explanations have been proposed: the replicability crisis in biomedical research, the so-called the file-drawer effect (5), misguided trust on model organism findings (6), unanticipated side effects (7), the implementation and management of clinical trials (8), excessive focus on a few genes (9), and so on. It’s important to note that most of these issues are not specific to drug discovery, but also occur in other domains of biomedical research.

Indeed, some researchers hold a pessimistic view of the usefulness of the primary scientific literature on informing translational research (10-12). In recent years, and in response to this disbelief, mounting efforts have been made to integrate target-disease evidence from diverse sources, with a special focus on large-scale, sequencing-based studies such as genome-wide association (GWAS) studies, transcriptomics, and massively parallel CRISPR-based genetic perturbation screening (13). The premise is simple: these large-scale studies provide massively more information per assay, are less impacted by human cognitive bias, and are often directly conducted on human cells or in living humans (14, 15). However, these large-scale studies tend to produce a lot of potential targets, among which only a few will be effective in clinical trials. It remains necessary for the scientific community to conduct careful downstream validation studies, with published literature as the main output. Thus, it is still important to ask whether scientists are able to align their research attention with respect to the translational potential of targets.

Here, we systematically analyze the evidence reported in clinical trial registration via the Open Targets platform (16), a database that integrates evidence from multiple sources, including genome-wide approaches, and biomedical literature. Open Targets is one of the most comprehensive harmonized public-data resources on interventional drug clinical trials (16). We find that clinical trials still predominantly rely on evidence that only exists in the primary research literature text, where small-scale, non-genome-wide studies are the norm (17). Furthermore, Pharma sponsors, particularly large pharmaceutical firms, appear to rely even more heavily on literature-grounded evidence than Non-pharma sponsors. To facilitate further systematic studies of the factors affecting drug development outcomes, we report here the first automated open pipeline to classify the success of individual clinical trials. Although trial information is openly available in the public domain (e.g., clinicaltrials.gov), it often lacks clarity on whether a trial is successful or not due to unstructured data. Our pipeline addresses this by leveraging the progression of trials through different phases— classifying only those advancing to the next phase as successes. This classification provides crucial insights that are not readily apparent from the raw data. We then use the pipeline to demonstrate that the intensity of disease-specific research on individual gene-disease pairings informs the success of drug clinical trials, while such alignment does not extend to non-disease research on human genes. These results have implications for scientific resource re-allocation in biomedical research. We also make our classified trials openly available to guide future research in this area.

### Data

We obtained the complete Open Targets database (release 24.09) from EMBL-EBI’s FTP site on 12/31, 2025. Open Targets incorporates harmonized disease-target evidence, FDA approved drugs with their initial approval dates, and curated trials data from ChEMBL (18). The ChEMBL database annotates drug clinical trials from clinicaltrials.gov using a custom pipeline, further enhanced by manual curation. The harmonized ChEMBL dataset in Open Targets encompasses **71,869** annotated drug clinical trials spanning different phases, including a small number of trials with ambiguous phase information.

In addition to these data, we sourced launch date, phases, and sponsor details for each trial from the AACT website (https://aact.ctti-clinicaltrials.org/download, version 20251231) on 12/31, 2025, and matched it with the curated trials in Open Targets using the ‘nct’ identifier. We were able to match 99% of the **83,744** trials. Based on the matched sponsors, these trials can be separated into Pharma or Non-Pharma led trials. We further breakdown the Pharma led trials into those led by Top-10 Pharma or other Pharma based on the total number of phases 2–4 trials launched during 2008-2019.

We focus our analysis on trials initiated between 2008 and 2019, although the underlying databases include releases through 2024-09. We chose 2019 as the endpoint for several reasons. First, ending the study period in 2019 leaves sufficient time for downstream trial linkage, which is essential for our automated pipeline for classifying trial success and failure. Second, trials initiated after 2019 may be affected by the COVID-19 pandemic, potentially confounding comparisons with earlier periods. An additional practical advantage of using 2019 as the endpoint is that it allows us to divide the study window into three equal-length periods: 2008–2011, 2012–2015, and 2016–2019, facilitating transparent temporal comparisons across phases of drug development. For completeness, we also provide basic descriptive statistics for trials initiated during 2020–2024 in Fig. S1.

### Linking disease-target evidence to clinical trials

The Open Targets platform integrates disease-target associations from 22 sources in addition to clinical trials, using standardized terminology. Each gene is identified by its Ensembl Gene ID, while diseases are classified using a hierarchically structured ontology system. This allows us to efficiently assess whether a disease-target pairing from a clinical trial is supported by other sources of evidence. We further organize the 22 sources of evidence into three major categories: genome-wide evidence, expert-curated evidence, and primary literature evidence.

Genome-wide evidence comes from genome-wide studies integrated into platforms like GWAS Catalog (19), EMBL-EBI Expression Atlas (20), and CRISPRbrain (21). Expert-curated evidence is manually selected by experts from clinical cases or literature. Primary literature evidence is derived from text-mined disease-target co-occurrences from EuroPMC (similar to PubMed) (22). Notably, when aggregated, primary literature evidence reflects the collective attention of the entire scientific community, unlike the summary statistics of genome-wide studies or the opinions of a few experts.

Additionally, Open Targets records metadata about the associated publications for each disease-target link in 15 out of 22 sources (16). This allows us to track the timing of target-disease evidence availability. Only evidence available before the launch of a clinical trial is considered supportive for that trial.

Beyond direct disease-target associations, Open Targets also infers indirect associations by propagating disease-target links downstream along the disease hierarchy. For instance, a target linked to ‘breast disorders’ would also be associated with its subcategories, such as ‘breast neoplasm’, ‘breast cancer’, and so forth. Due to the difficulty in quantifying the strength of such indirect evidence, we restricted our analysis to direct disease-target links.

## Results

### Open Targets data reveal the changing landscape of drug clinical trials

Figure 1a displays the number of newly initiated drug clinical trials cross time, segmented by phase. Since the FDA mandated the registration of all interventional phases 2–4 trials in 2007 (23), we focus on post-2007 data to capture the overarching trends in trial activity more accurately. As one would expect, the progression from one phase to the next in clinical trials is contingent on the success of the preceding phase. Therefore, we should observe a higher number of phase 2 trials than phase 3 trials and, likewise, more phase 3 trials than phase 4 trials. This pattern is indeed reflected in our data (Fig. 1a), confirming the conditional nature of the clinical trial progression. Interestingly, despite these expectations, phase 1 trials, which are the initial phase, do not account for the highest number of trials (Fig. 1a). This discrepancy suggests an under-reporting of phase 1 trials. Consequently, our analyses primarily concern phases 2-4 trials.

**Fig. 1.**
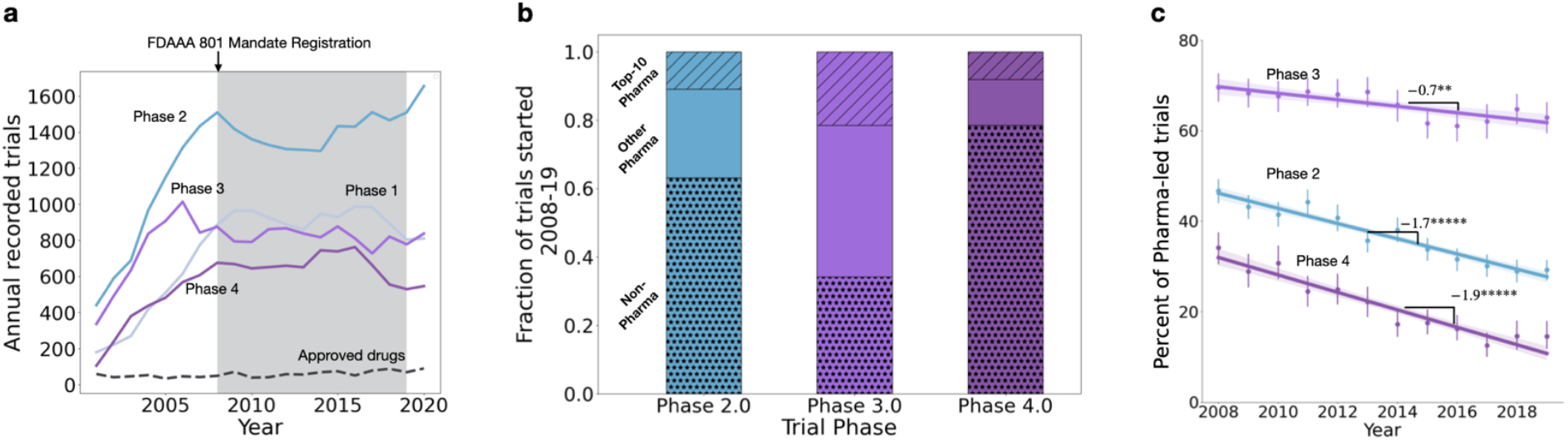
Data from clinicaltrial.gov suggests that a reporting steady-state exists for the period 2008-2019 and that the role of the Pharma industry in clinical trials has been systematically decreasing. **a)** Annual number of clinical trials broken down by phase, with the shaded area indicating the time frame studied (**2008-2019**). The clinicalgtrials.gov site was opened in Feb 2000, and a mandate for registration of phases 2-4 trials was issued in Sep 2007. This led to essentially full registration by 2008, for all but Phase 1 trials. **b)** Fraction of trials led by various sponsors during the **2008-2019** timeframe, broken down by phase and sponsor. Surprisingly, Pharma dominates Phase 3 trials but not other Phases. The Pharma sector is further breakdown into Top-10 and other Pharma based on the total number of phases 2-4 trials launched during **2008-2019. c)** Time evolution of the proportion of trials led by Pharma companies, broken down by phase. It is visually apparent that the fraction of Pharma led trials has been steadily decreasing. Number reports slope estimate and stars report statistical significance (**: p < 0.01; *****: p < 10^-5^; Pearson’s correlation test).

Despite the relatively high and stable number of trials across phases during our focus period, the number of newly approved drugs remains remarkably low, indicating a challenging landscape for drug development success (Fig. 1a). Despite the stability of the aggregate numbers, there are intriguing shifts in trial characteristics across phases and over time. For example, while pharmaceutical companies are instrumental in phase 3 trials, other types of organizations lead the charge in phases 2 and 4 trials (Fig. 1b). Moreover, in line with previous studies (24), the proportion of trials led by pharmaceutical companies, irrespective of the trial phase, has been on a downward trajectory (Fig.1c; p < 0.01 for all correlations plotted). This trend thus suggests a shift in the landscape of clinical developments, with Non-pharma sponsors becoming increasingly instrumental.

### Primary research literature is the most common source of evidence for drug clinical trials

As mentioned in the “Linking disease-target evidence to clinical trials” section, we use the Open Targets platform to identify the main source of support for drug clinical trials. In total, we identified support for ∼70 % of all trials. As can be seen in Fig. 2a, the largest source of such evidence is the primary literature (EuroPMC) (22).

**Fig. 2.**
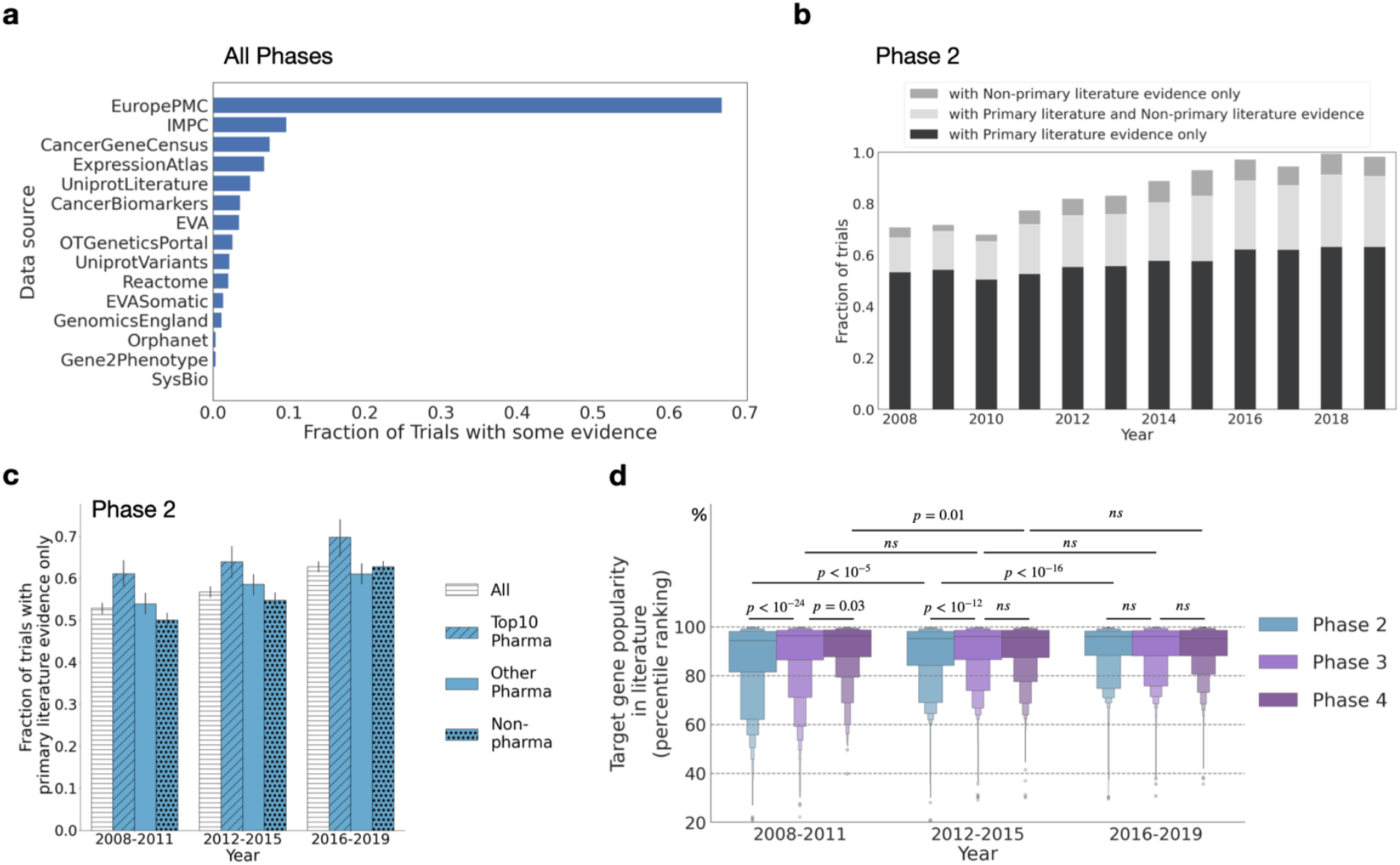
Primary literature evidence plays a dominant role in launching trials. **a)** Fraction of clinical trials (all phases) with evidence by source. The largest source of evidence is from primary literature text (EuropePMC). **b**) Fraction of phase 2 trials with some evidence supported increases across years. However, the dominant type of evidence is still primary literature text evidence. **c)** Pharma, especially Top-10 Pharmaceutical companies, relies more on text-mined primary literature evidence. **d)** Trials at all phases focus on highly popular genes. Interestingly, Phase 3 trials tend to focus on genes with a higher popularity than phase 2 trials. Moreover, the popularity of genes in phase 2 trials increases across years. For each trial, we compute the average popularity rank of the target genes and plot the letter value plot of these averages across all trials, breakdown by phases and periods. P-values are computed using Mann–Whitney U tests.

Sources from EuroPMC are used to justify at least 65% of annotated trials (Fig. 2a). The observation that the primary literature is the most common type of evidence still holds true after attributing trials supported by both the literature and some other annotation source to the latter (Fig. 2b). This pattern is consistent across all trial phases (Fig. S2). Further, Pharma sponsors, particularly the Top-10 Pharma sponsors, appear to rely even more strongly on literature-based evidence than Non-pharma sponsors (Fig. 2c). This pattern further underscores the importance of the primary research literature in rationalizing the initiation of clinical trials, even though the reliability of such evidence has been questioned by many stakeholders (10-12).

A question prompted by these observations is: *how* do trial sponsors select which evidence from the literature surrounding potential targets to focus on? Prior research has demonstrated that most literature is focused on a small fraction of genes (25). To investigate whether those highly popular genes are also the focus of literature-based evidence in drug clinical trials, we obtain from Open Targets the number of publications reporting studies on each human protein-coding gene prior to the initiation of the trial (see Methods for details). We observe that identified drug trial target genes tend to be selected from among the small set of highly studied genes (Fig. 2d, see Methods for definition of target popularity in a trial). Furthermore, we observe that the median popularity of the genes targeted increases over time for phase 2 trials and from phase 2 to higher phases. As we only consider literature preceding the trials, accumulation of literature on a gene appears essential for relevant drug development.

In summary, the findings reported in Fig 2 highlight the importance of primary literature evidence to the initiation of drug trials. However, this finding does not provide insights into the effectiveness of primary literature evidence on trial success.

### Developing and validating an open pipeline for assessing clinical trial success

To test if the abundance of primary literature can inform on the success of clinical trials, we need to first discriminate between successful and unsuccessful interventions against a given disease. In principle, the information on the success of trials should be public. In practice, however, there is no public resource that connects distinct trials and thus provides the progress of interventions for all publicly accessible trials. To address this need we developed an automated open pipeline based on public information gathered and harmonized by Open Targets (Fig. 3a; see also Methods and Fig. S3 for a detailed breakdown for phase 2 and phase 3 trials separately). To promote further research, we make the dataset created by this pipeline accessible as Supplemental Table 1.

**Fig. 3.**
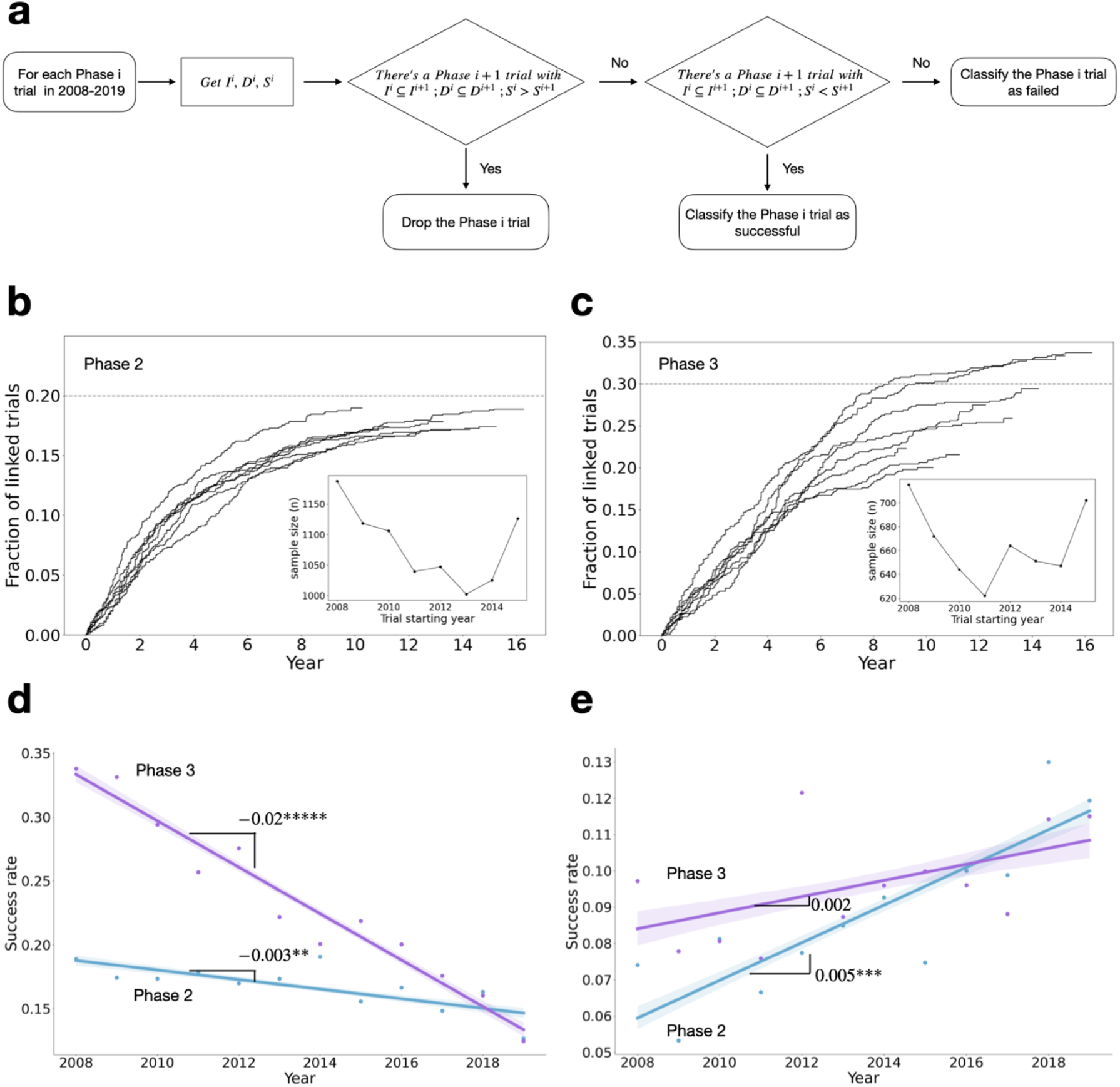
**a)** Open pipeline for following the success of drug development along consecutive clinical trial stages. Because different Phases of the trial for a specific drug are not linked in the data, we developed a protocol for linking them. For a Phase i trial (T^i^), we have information on start date (S^i^), set of drugs studied (D^i^) and set of indications for those drugs (I^i^). A Phase i+1 trial (T^i+1^) can be linked to T^i^ if and only if S^i+1^ > S^i^, all I^i^ in I^i+1^, and all D^i^ in D^i+1^. If a trial T^i^ is linked to at least one Phase ^i+1^ trial, then it is deemed to be successful. Trials without any higher Phase linkages are deemed unsuccessful. For two linked trials, we denote S^i+1^-S^i^ as the time gap between linked trials. **b)** Fraction of linked phase 2 trials launched during 2008-15. The trend of linked probabilities is similar across years, and plateaus at around 20%, corresponding to an ultimate success rate of around 20%, consistent with the value estimated by the FDA (25-30%). **c)** Fraction of linked phase 3 trials launched during 2008-15. The trend for fractions linked is similar across years, and plateau at around 30%, corresponding to an ultimate success rate of around 30%, consistent with the value estimated by the FDA (33%). **d)** Success rate by launch year. If no time horizon is placed on estimation of success rates, then it would appear that trials—especially Phase 3 trials—become less successful over time. **e)** Success rate by launch year with a 5-year time horizon. When a 5-year time horizon is considered when calculating the success rate, the apparent trend of decreasing successful rate reversed. The numbers report slop estimates and the stars report statistical significance for Pearson’s correlation test (*****: p < 10-5; ***: p < 0.001; **: p < 0.01).

Our pipeline categorizes a phase *i* trial as successful if it is associated with at least one subsequent phase *i*+1 trial. All trials for which no such association is found are categorized as unsuccessful. We further categorize a trial as ΔT successful if an association can be established within T years of the start of the trial. We can assign a success/failure categorization to 13,479 phase 2 (73% of total), and to 7,936 phase 3 trials (71.5% of total). Our trial categorization pipeline has an overall agreement rate of 84% with the categorizations of a previously published, smaller scale (983 trials), semi-manually curated dataset (26) (Fig. S4).

To validate the quality of our trial linkages, we first estimated the overall trial success rate for both phase 2 and phase 3 trials using the Kaplan-Meier model (refer to the section “Estimating the overall trial success rate” in Methods for more information). As depicted in Fig. 3b, for phase 2 trials, the trend for the fractions of linked trials remains consistent over time, eventually stabilizing at roughly 20%. This figure corresponds to a final success rate of around 20%, mirroring the FDA’s estimate of 25-30%. For phase 3 trials, we determined a relatively steady success rate of 30% (Fig. 3c), which is again in line with the FDA’s estimate of 33%.

Due to the granularity of our data, we can investigate whether there are any temporal trends in the success rate of clinical success. Fig 3 b and c suggest that it is important to adjust for data censoring. If one does not adjust for data censoring, then one would conclude that trials success rates have been decreasing over time (Fig. 3d). However, as shown in Fig. 3e, when considering the success rate within a fixed time window (5 years after the start of a lower-phase trial), we find an increasing success rate over time.

### Characteristics of literature-reported evidence inform on the success rate of drug clinical trials

We are now able to assess whether there are associations between the characteristics of prior primary research literature selected as evidence for a drug clinical trial and its success. As the results of genome-wide approaches can yield subsequent literature, we seek to improve interpretability by excluding trials that besides literature also carry any other source of support in Open Targets.

To quantify the characteristics of the literature-based support selected for supporting the trial, we calculate the average popularity of all disease-target associations reported for that trial using literature published before each trial’s starting year. Similar to Fig. 2, we transform the raw publication counts into percentiles as the total volume of literature grows over time. As demonstrated in Figs. 4a and b, successful trials are associated with higher disease-target popularity, a pattern consistent across phases and time periods (p < 0.009 for all comparisons). This association is strengthening over time and is particularly strong for phase 3 trials. Note that similar conclusions can be drawn if one defines the characteristics of the literature-based support of a trial by calculating the median of the popularities within the trial instead of the mean (Fig. S5 a,b). Similar patterns were observed when differentiating between Pharma-led and Non-pharma led trials (Fig. S6), despite the differences becoming less statistically significant in some time periods, likely due to the lack of statistical power.

**Fig. 4.**
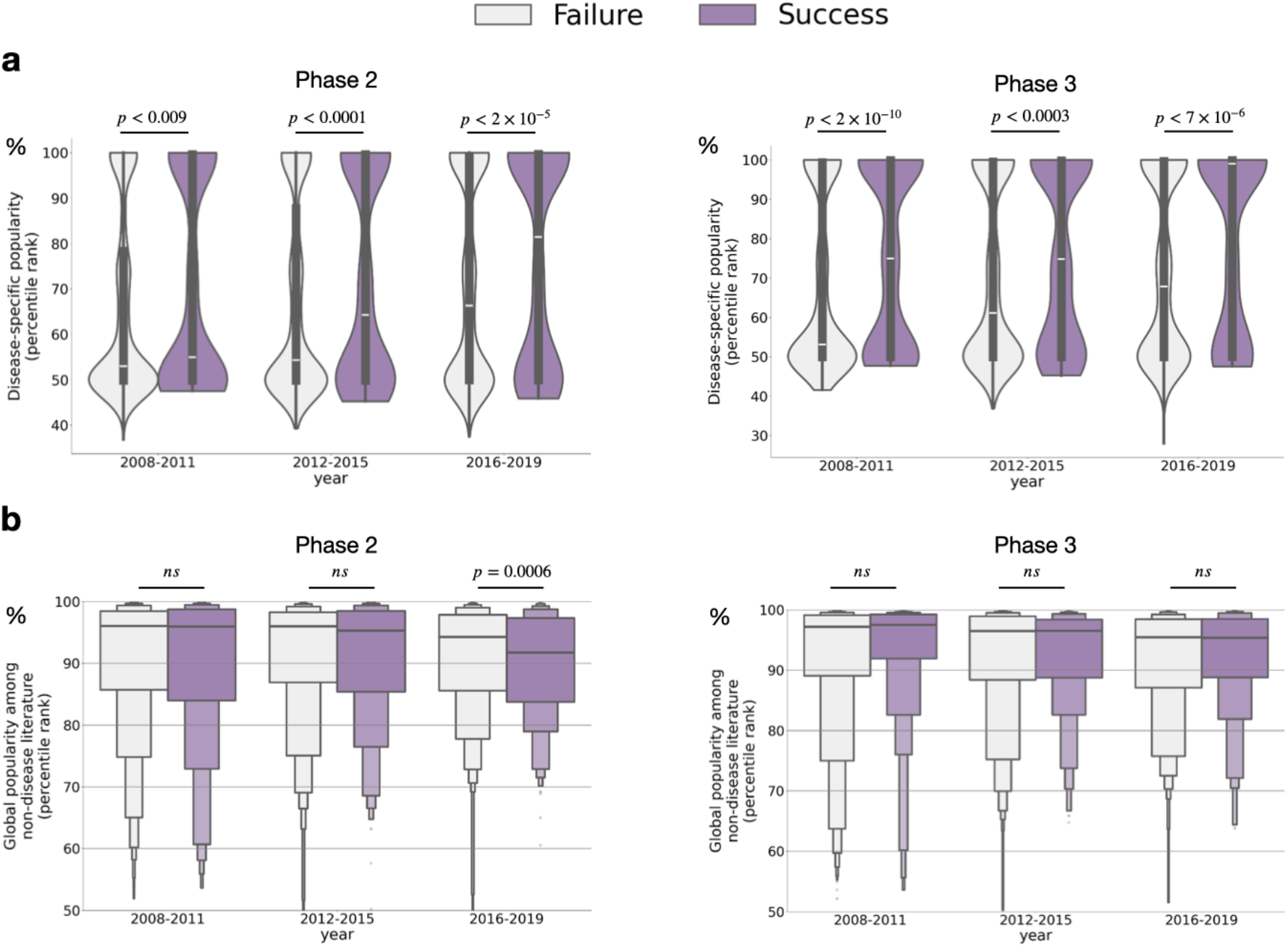
Disease-specific target popularity, but not global target popularity from non-disease human gene research, is predictive for clinical success. For each trial, we compute the average popularity rank of the disease-target combinations (or targets) and plot these averages across all trials, breakdown by phases and periods. **a)** Disease-specific popularity is significantly higher in successful Phase 2 trials versus failed Phase 2 trials and successful Phase 3 trials versus failed Phase 3 trials. **b)** Global target popularity from non-disease research in successful phase 2 trials is not significantly higher compared to failed phase 2 trials (during 2016–2019, the popularity of target genes was even significantly lower than that of failed trials), nor for the comparison of successful and failed Phase 3 trials. All p-values are computed using two-sided Mann-Whitney U test.

The above findings demonstrate the alignment between collective attention in disease-specific research and clinical trial success. One might wonder whether such alignment extends to more basic research, such as studies on human genes without specific disease relevance. On one hand, as suggested by Fig. 2d, only genes that are popular in literature tend to enter clinical trials. On the other hand, predicting the disease relevance of a gene is challenging, and in non-disease research, scientists might not choose genes based on disease relevance. To explore this, we first estimate the average global popularity of genes in trials, similar to our approach in Fig. 2d, but focusing on literature that does not mention any disease. We then compare this popularity between successful and failed trials. As shown in Fig. 4c and d, the attention given to genes in non-disease research is *not associated* with clinical success. Using the median instead of the mean for aggregating popularity within a trial yields similar results (Fig. S5 c,d).

Overall, our findings suggest a highly context-dependent alignment between scientific research and clinical success.

### Limitations

A limitation of the data we analyze is that they do not provide information about evidence used to rationalize roughly 20-30% of clinical trials. It is extraordinarily unlikely that a trial would be initiated without some form of evidence supporting it. We suspect this void may be partly due to incomplete literature annotation, as our findings were exclusively sourced from open data, potentially excluding evidence behind paywalls or classified as trade secrets. Additionally, some drugs in clinical trials, like Aspirin, are identified by their physiological effects without knowledge of their cellular targets (27). The dominant mechanism behind this “missing evidence” puzzle thus remains to be explored.

One potential confounding factor when quantifying the aggregate popularity of the disease target in the literature-based evidence is the inclusion of targets from control drug interventions in addition to the targets of new interventions to be tested. Within a trial, aggregating the disease target popularity by the mean, or even the median, might not accurately reflect this fact. To test if this caveat may affect our conclusions, we also investigate the same hypothesis using the minimum popularity of all disease-target associations reported for that trial, as new interventions are more likely to involve newer, less popular genes than old interventions. Reassuringly, we recover the conclusions supported by Fig. 4 under this extreme choice (Fig. S7).

Finally, our work could be extended along multiple directions. First, while we identified a difference in evidential characteristics between successful and failed trials, the effect size is modest. This may be due to our reliance on the simple co-occurrence frequency of disease-target pairs, a method that cannot account for nuanced context that would require a full textual understanding. The increased sophistication of large-language models may yield more precise aggregation methods for such textual evidence (28). We expect the effect size to increase when accounting for reasons of clinical trial failures that are beyond target selection (3,8), most notably, insufficient patient enrollment (8). Second, we only study target-disease linkages that enter clinical trials. However, many reported target-disease linkages do not enter clinical trials. Are these un-examined linkages known to be irrelevant, or are they neglected potential innovations?

## Discussion

We examined the role of primary literature evidence sources in shaping drug clinical trials by integrating and analyzing the information contained in Open Targets, a public repository of clinical trials and disease targets. We found that the primary scientific literature remains the most important evidence source in recent trials. In addition, we demonstrated that only genes that have been the focus of a significant degree of research attention—i.e., popular genes—will be likely to be considered in the selection of evidence for supporting a drug clinical trial (Fig. 2c). A recent study on network effects on drug discovery (29) suggests that this may be due to preferential attachment — i.e., copying behavior — in drug-target selection.

By developing an open pipeline to infer success rates of clinical trials, we can ask whether the prior research attention can predict clinical success. We found that disease-specific attention toward a gene indeed has some predictive power for clinical success. However, our data also reveal that across all literature in Open Targets, basic research attention *is not* associated with greater success rates for drug clinical trials in which they are identified as targets (Figs. 4c and d). This raises the interesting question of whether there would be ways of further improving the overall efficiency of basic research. Our open pipeline is a starting point for these efforts, providing accessible clinical translation data to scientists who may be unaware of how their basic research impacts real-life applications.

While our data suggests that the primary research literature remains the principal source of evidence for initiating clinical trials, it’s important to consider the relatively recent emergence of disease-target relationship based on genome-wide evidence (30). At present it is not clear whether detailed study of selected disease-target relationships presented in the form of primary literature will maintain a key role in guiding clinical trials or whether data-driven methods may simply need time to gain momentum and surpass prior primary research approaches where genome-wide evidence is not the major source. Despite the uncertainty, our analysis suggests that the proportion of trials supported by primary literature has been increasing over time when considering evidence presented simultaneously in both primary literature text and other sources (Fig. 2b). This trend underscores the growing potential of primary research to effectively integrate data-driven evidence and remain essential in the future.

We anticipate that the open data on the success of clinical trials, which we presented in this manuscript, will spur further research into the success and failure of clinical trials, but also into the reverse problem—how the outcomes of clinical trials may impact the direction of fundamental research (31). We hope our study will stimulate more effort into building a two-way street between the bench and the bedside.

## Methods

### Bibliometric data

The Open Targets text-mining data annotate mentioning of biomedical entities, including genes, species, disease for individual papers from EuroPMC (equivalent to PubMed) sentence by sentence. In our analysis below, we define a paper mentioning an entity as long as the entity is mentioned at least once in any section of the scientific literature catalogued by EuropePMC.

### Target gene popularity

For each target gene listed in a target-disease pair in a trial, we calculate the number of unique publications that refer to that gene up to the starting year of the trial. After we calculate this number for every protein-coding human gene, we rank their values and determine a percentile score. A higher percentile rank indicates a greater popularity for the target. The target popularity among non-disease research is computed similarly among literature that did not mention any diseases.

When considering target-disease pairs, we follow the definition of disease-target linkage in the Open Targets platform. Briefly speaking, Open Targets consider a disease-target has literature support as those disease-target pairs that are mentioned in the same sentence in at least one literature. For each disease-target pair, we count the number of publications that co-mentioned the disease and target in at least one sentence in any section. For disease-target pair without any co-mentions, we consider the count as zero. Subsequently, we rank the disease-target count within each disease. Because each ranking is computed for a specific disease, these rankings can be substantially different from the global target popularity ranking. Indeed, as of 2019, the median Pearson correlation between disease-specific target popularity and global target popularity is only 0.07.

### Evidential strength supporting a trial from literature

The evidential strength supporting a trial is computed from the popularity of targets within the relevant literature published before each trial’s commencement. This popularity is determined by counting the occurrence of each gene in literature related to a specific disease (assigning a count of zero to genes not mentioned before the trial launch). Subsequently, we standardized this count to a value between zero and one through percentile ranking. Finally, we quantify the evidence supporting each trial by calculating the mean, median or min ranking of all disease-target associations within that trial.

### Classifying the outcome of a trial

We start by assigning a unique phase to each trial. Note that besides the standard phases 2, 3, and 4, some trials are also designated as phase 1/2 or phase 2/3. Following the practice in the literature (32) we denoted 3051 phase 1/2 trials as phases 2 and 1332 phase 2/3 trials as phase 3.

We categorize a phase i (= 2,3) trial as successful if there was at least one subsequent phase i+1 trial encompassing all indications (i.e., diseases) and drugs from the phase i trial. During this process, we noted 8,142 instances (4,979 phase 2 trials and 3,163 phase 3 trials) where a phase i trial was linked to one or more phase i+1 trials that launched before the start of the phase i trial. For these trials, their chances of linking to some i+1 phase trials in the future are also very high (76% for phase 2 trials, and 75% for phase 3 trials). There are two potential possibilities of the high rate of linkage. One possibility is that, interventions that were successful in the past also tend to be successful if tested again, presumably for further optimizing doses. However, another possibility is that these are simply false positives: some key interventions in a trial are missing, and the matched interventions are all placebo interventions which tend to be standard comparisons in different trials. To see what the main driving force of the high linkage rate is, we manually reviewed 20 randomly sampled instances from those 8,142 trials and found that 50% were false positives. These false positives resulted from unannotated drugs in the phase i trials, thereby causing incorrect linkages. Conversely, an examination of 20 randomly-sample phase i+1 trials that could not be connected to any preceding phase i trial showed that only 20% had omitted drug annotations. Informed by these observations, we excluded trials that were linked to later phase trials but that had start dates earlier than the start date of the focal trial. In total, our procedure yielded the classification of 13,479 phases 2, and 7,936 phase 3 trials starting in 2008-2019.

### Overall trial success rate

To estimate the overall success rate, we leverage the event status (linked or unlinked) and the associated time-to-event for each trial. Specifically, for a linked phase *i* trial, the time-to-event is the interval between the inception of the trial and the initiation of the earliest subsequent phase i+1 trial. For an unlinked trial, the time-to-event is the span between the trial launch and the final recorded date of any phase i+1 trial in our database, known as the right censoring date. This data allows us to construct a Kaplan-Meier curve using standard methods. We produce separate Kaplan-Meier curves for trials initiated in each year from 2008 through 2015. We show the results for phase 2 and phase 3 trials in Figs. 3b and 3c, respectively.

## Data Availability

Data is already publicly available.

## SUPPLEMENTARY FIGURES

**Fig. S1.**
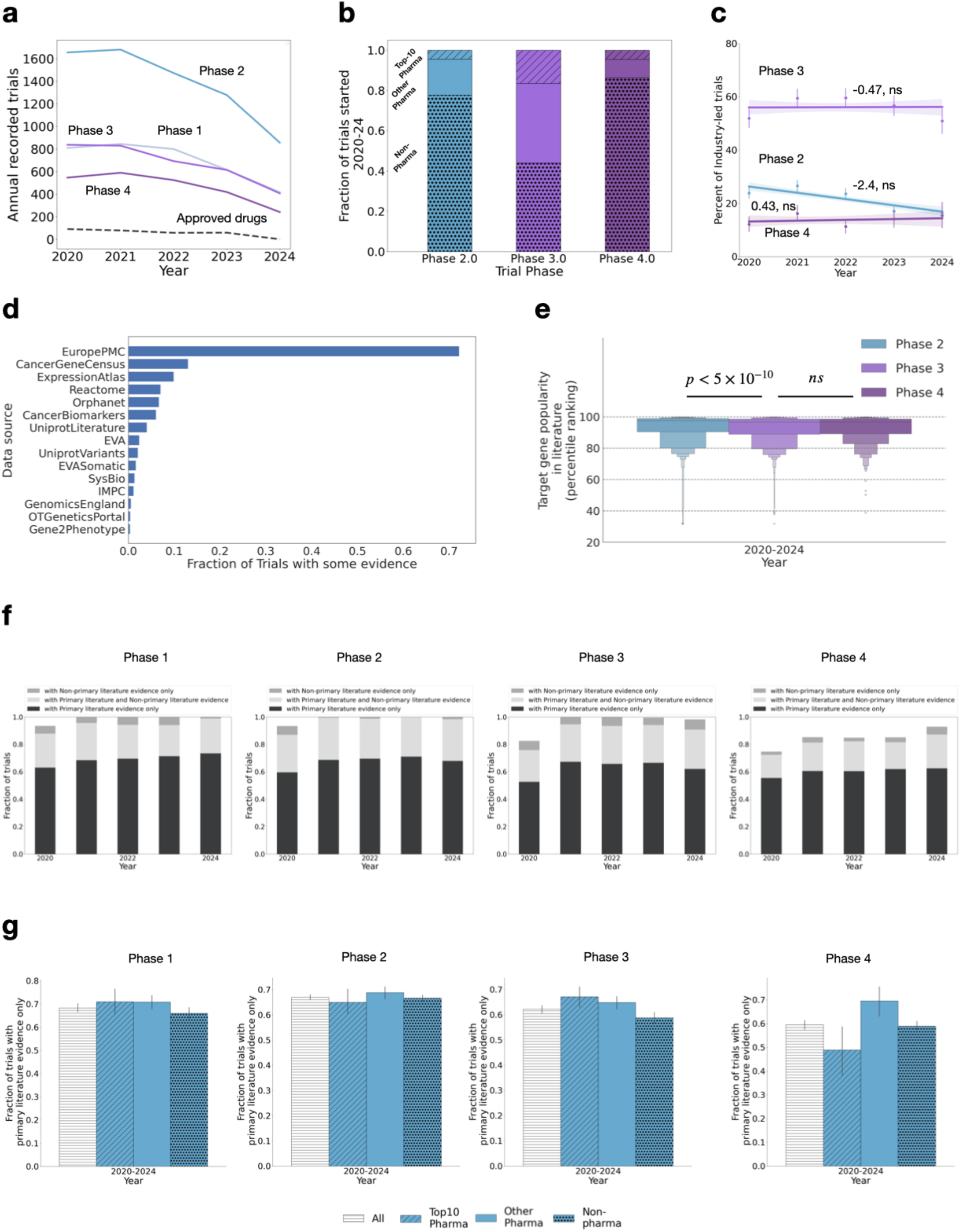
**a)** Annual number of clinical trials broken down by phase during **2020-2024. b)** Fraction of trials led by various sponsors during the **2020-2024** timeframe, broken down by phase and sponsor. **c)** Time evolution of the proportion of trials led by Pharma companies, broken down by phase during **2020-2024. d)** Fraction of clinical trials (all phases) with evidence by source during **2020-2024. e)** Trials at all phases focus on highly popular genes during **2020-2024. f)** Fraction of phase 1,2 ,3 and 4 trials with some evidence supported increases across years **2020-2024. g)** Reliance on Literature Evidence in Phase 1, 2, 3, and 4 Trials during **2020-2024**, by Sector.

**Fig. S2.**
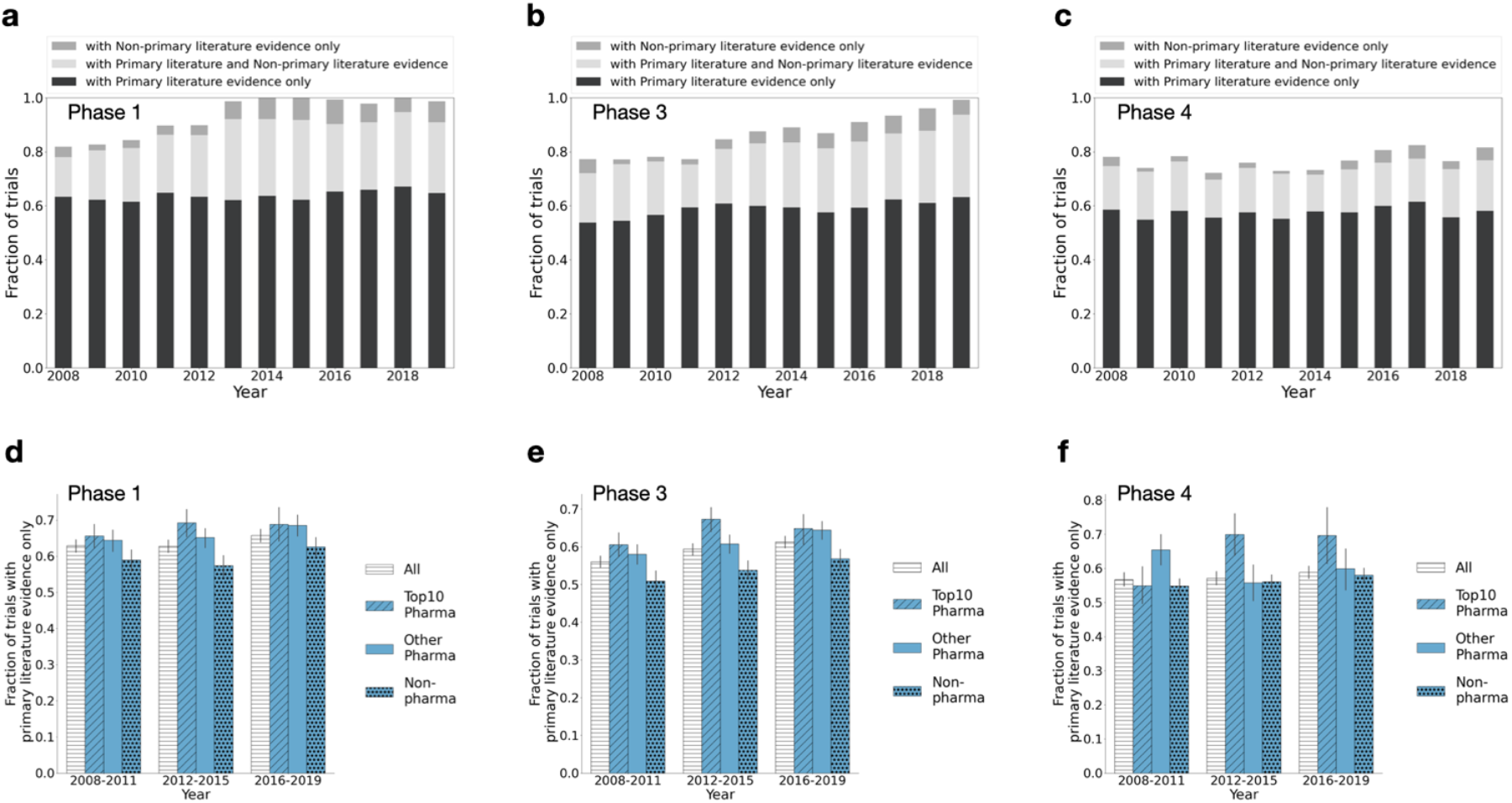
**a)-c)** Fraction of phase 1,3 and 4 trials with some evidence supported increases across years. However, the dominant type of evidence is the primary research literature. d**) - f)** For phase 1,3 and 4 trials, Pharma, especially Top-10 Pharma, relies more on the primary research literature as evidence.

**Fig. S3.**
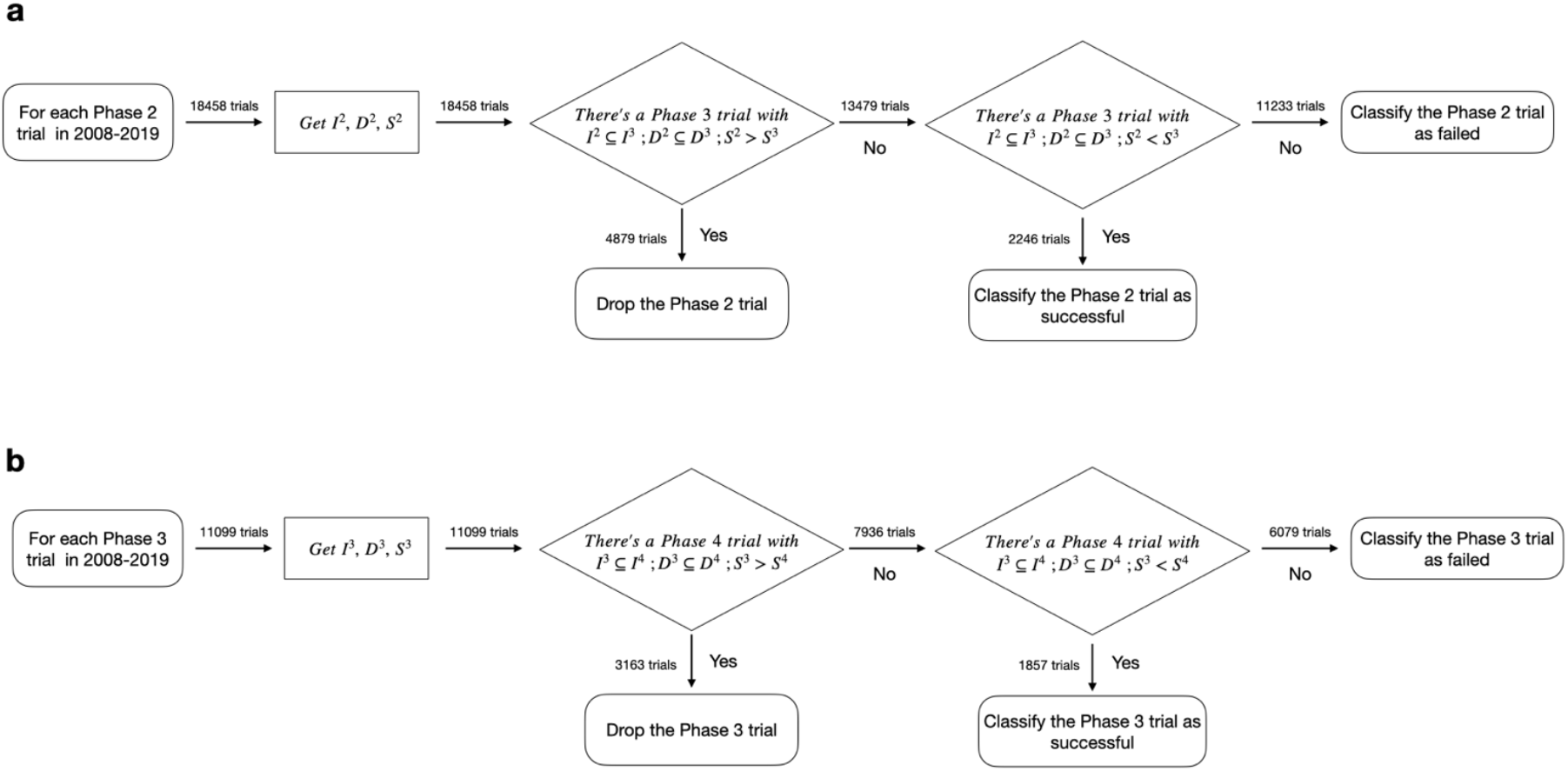
Classifying the outcome of trials. Same as Fig.3a but with detail statistics for (a) Phase 2 trials and (b) Phase 3 trials separately.

**Fig. S4.**
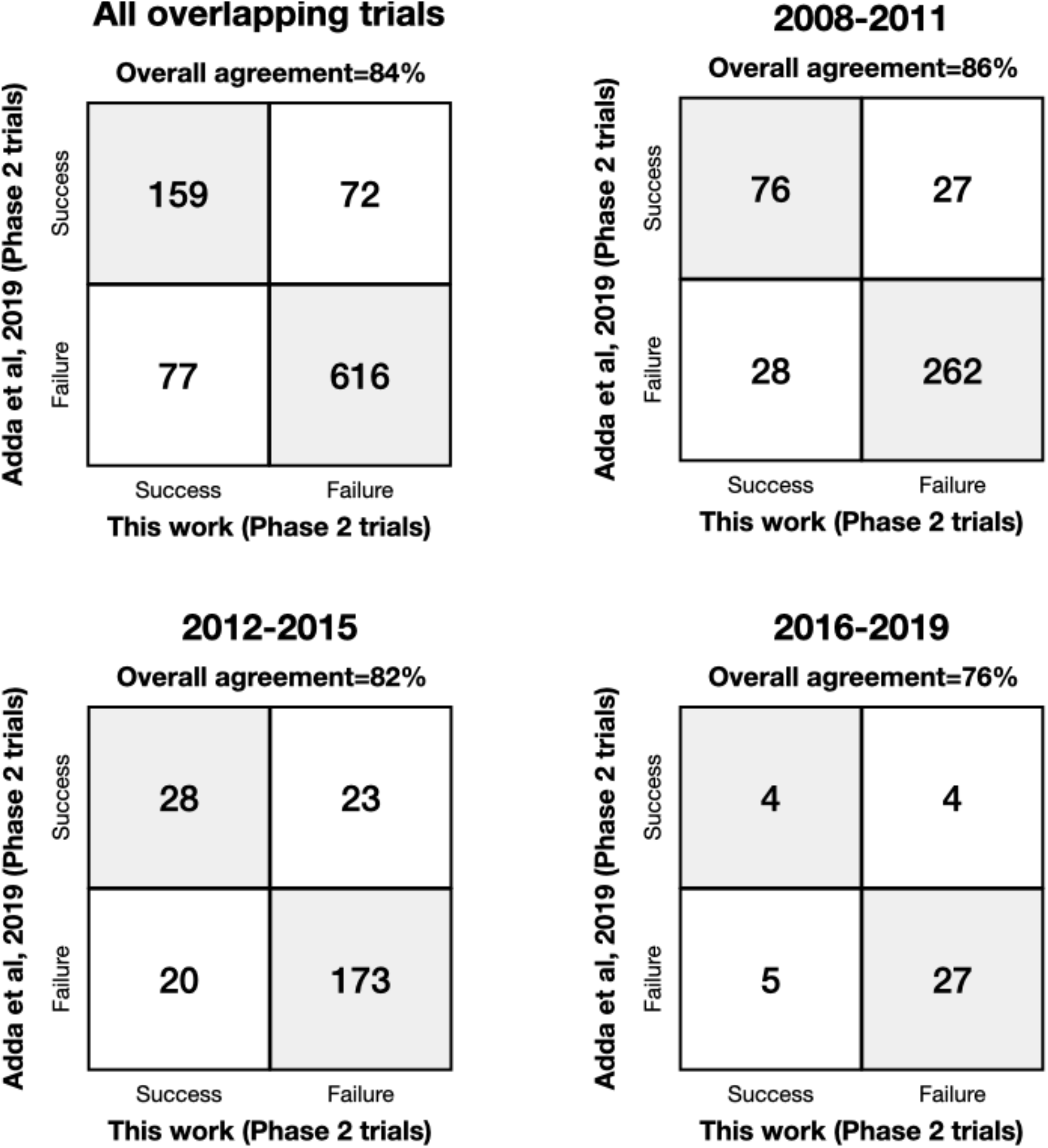
Comparison with a recent study that manually matched a small subset of Phase 2 trials. Our procedure yields highly concordant results for all overlapping trials (not limited to study period 2008-2019), and the concordance is similar across different time periods, supporting the reliability of our automated linking procedure.

**Fig. S5.**
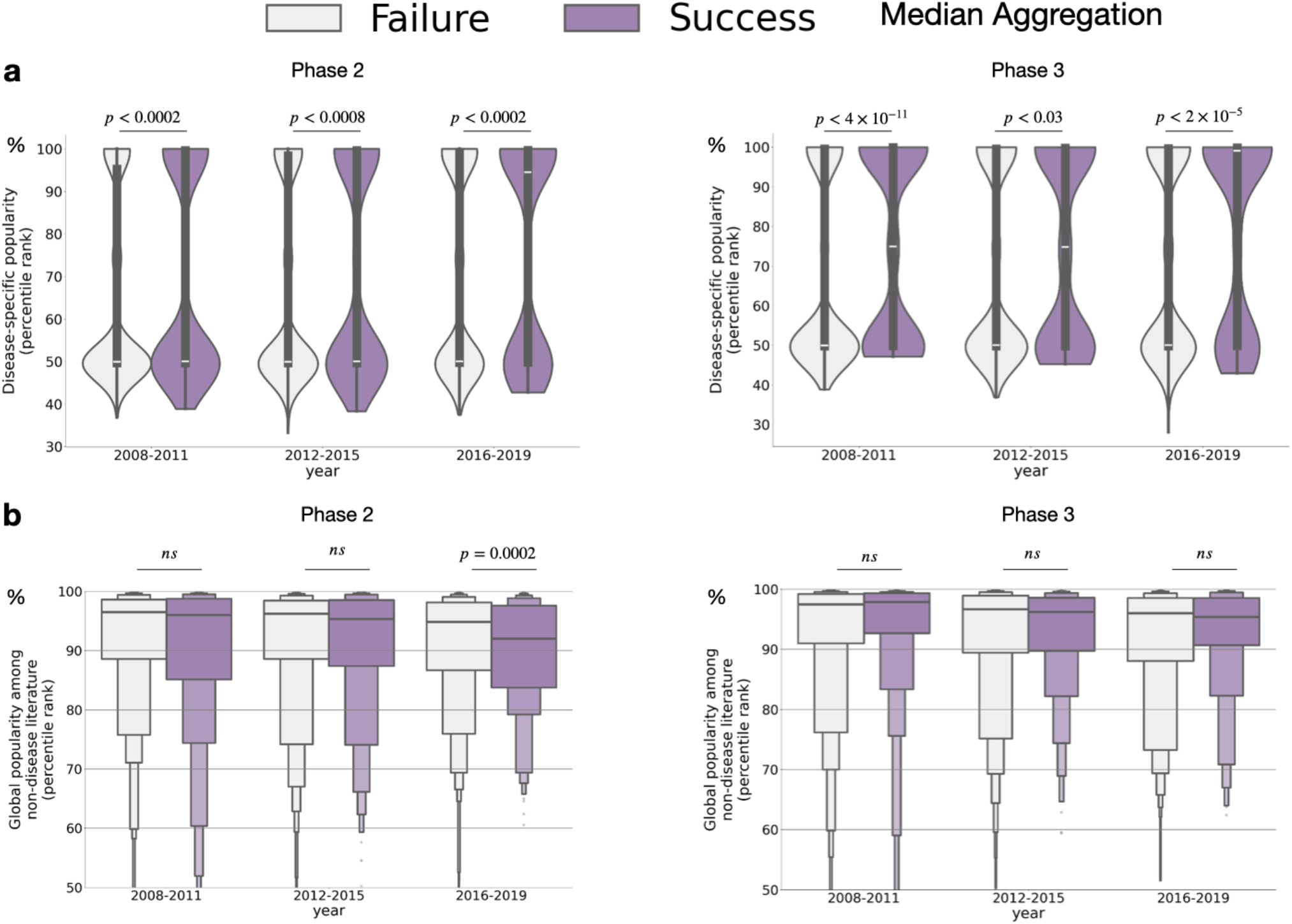
Same as Fig. 4 but using median instead of mean to aggregate target popularity of a trial.

**Fig. S6.**
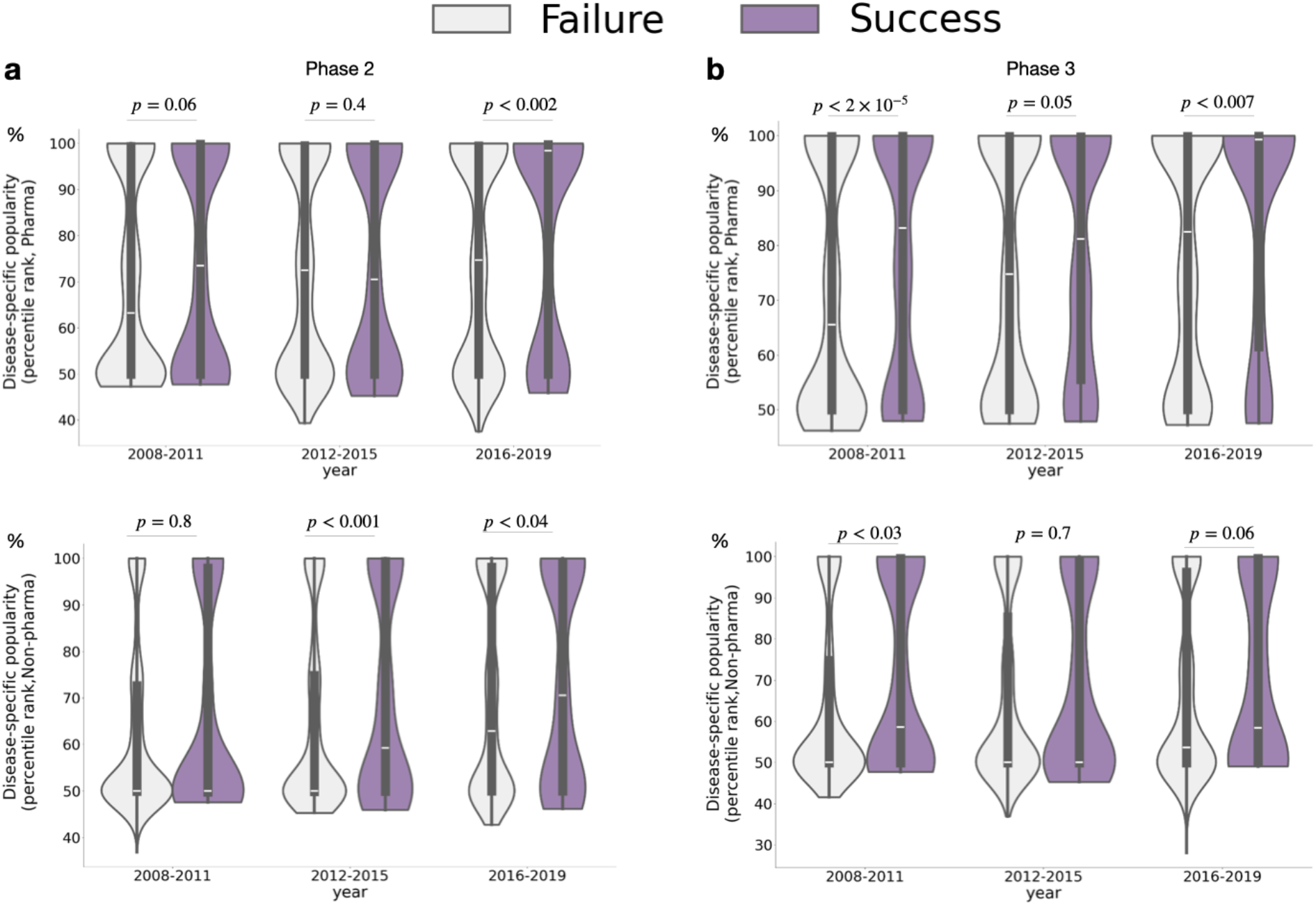
**a)** Comparison of disease-specific target popularity between successful and failed Phase 2 trials, breakdown by Pharma-led trials and Non-pharma led trials. **b)** Comparison of disease-specific target popularity between successful and failed Phase 3 trials, breakdown by Pharma-led trials and Non-pharma led trials. All p-values are computed from two-sided Mann-Whitney U test.

**Fig. S7.**
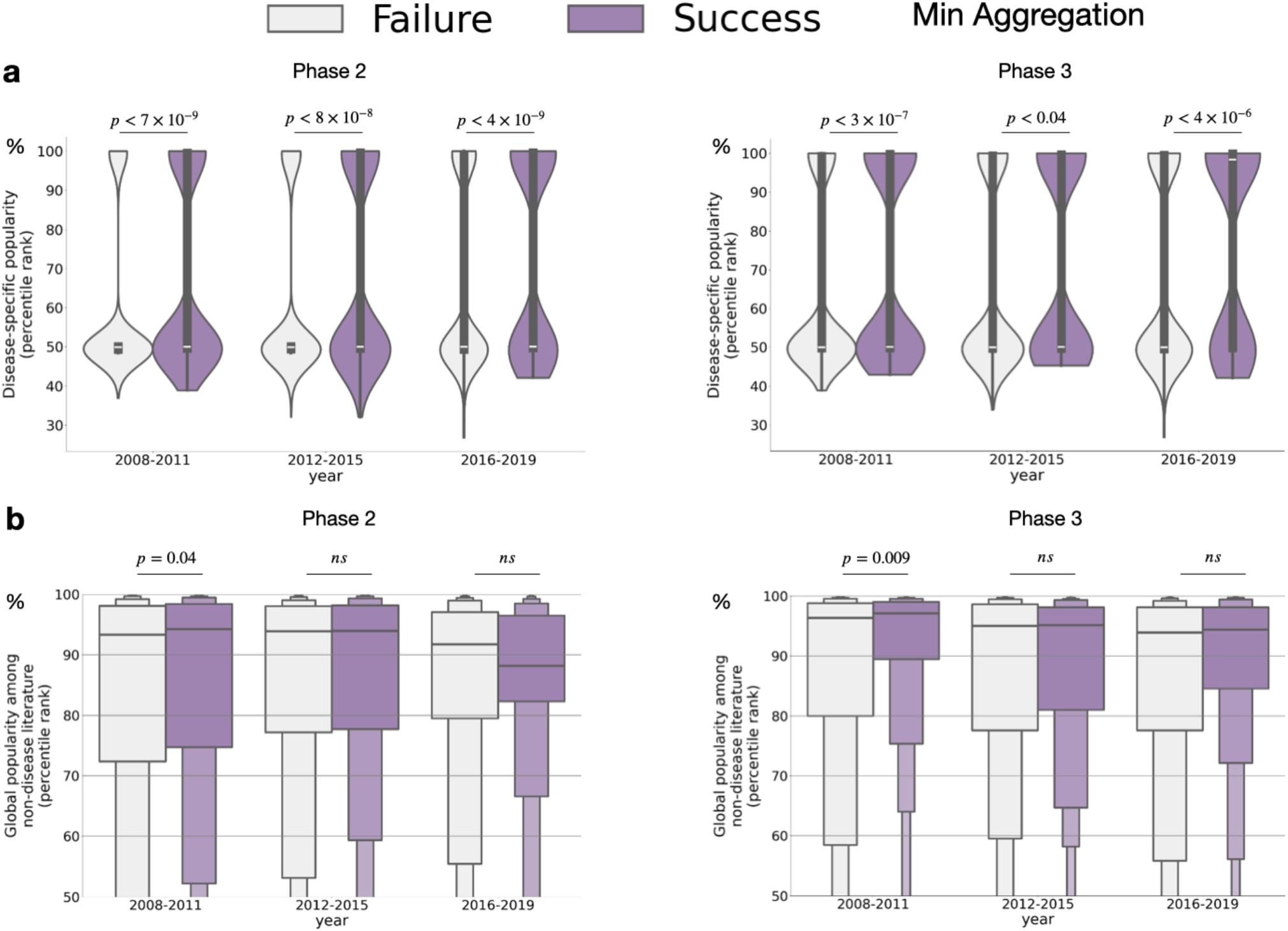
Same as Fig. 4 but using minimum instead of mean to aggregate target popularity of a trial.

## Notes

### Competing Interest Statement

The authors have declared no competing interest.

### Funding Statement

T.S. is supported by R00AG068544 and NSF 2410335

### Summary of Updates

Update figures with newer version of data

## References

1. Hughes, James P., et al. “Principles of early drug discovery.” British journal of pharmacology 162.6 (2011): 1239–1249.

2. Melsheimer, Richard, et al. “Remicade®(infliximab): 20 years of contributions to science and medicine.” Biologics: Targets and Therapy (2019): 139–178.

3. Harrison, Richard K. “Phase II and phase III failures: 2013–2015.” Nat Rev Drug Discov 15.12 (2016): 817–818.

4. Hay, Michael, et al. “Clinical development success rates for investigational drugs.” Nature biotechnology 32.1 (2014): 40–51.

5. Ioannidis, John PA. “Why most published research findings are false.” PLoS medicine 2.8 (2005): e124.

6. McGonigle, Paul, and Bruce Ruggeri. “Animal models of human disease: challenges in enabling translation.” Biochemical pharmacology 87.1 (2014): 162–171.

7. Moore, Thomas J., Bruce M. Psaty, and Curt D. Furberg. “Time to act on drug safety.” Jama 279.19 (1998): 1571–1573.

8. Nipp, Ryan D., Kessely Hong, and Electra D. Paskett. “Overcoming barriers to clinical trial enrollment.” American Society of Clinical Oncology Educational Book 39 (2019): 105–114.

9. Oprea, Tudor I., et al. “Unexplored therapeutic opportunities in the human genome.” Nature reviews Drug discovery 17.5 (2018): 317–332.

10. Dolgin, Elie. “Drug discoverers chart path to tackling data irreproducibility.” Nature Reviews. Drug Discovery 13.12 (2014): 875.

11. Prinz, Florian, Thomas Schlange, and Khusru Asadullah. “Believe it or not: how much can we rely on published data on potential drug targets?.” Nature reviews Drug discovery 10.9 (2011): 712–712.

12. Begley, C. Glenn, and Lee M. Ellis. “Raise standards for preclinical cancer research.” Nature 483.7391 (2012): 531–533.

13. Ghoussaini, Maya, et al. “Open Targets Genetics: systematic identification of trait-associated genes using large-scale genetics and functional genomics.” Nucleic acids research 49.D1 (2021): D1311–D1320.

14. Tam, Vivian, et al. “Benefits and limitations of genome-wide association studies.” Nature Reviews Genetics 20.8 (2019): 467–484.

15. Bock, Christoph, et al. “High-content CRISPR screening.” Nature Reviews Methods Primers 2.1 (2022): 1–23.

16. Ochoa, David, et al. “Open Targets Platform: supporting systematic drug–target identification and prioritisation.” Nucleic acids research 49.D1 (2021): D1302–D1310.

17. Stoeger, Thomas, and Luís A. Nunes Amaral. “The characteristics of early-stage research into human genes are substantially different from subsequent research.” PLoS biology 20.1 (2022): e3001520.

18. Gaulton, Anna, et al. “ChEMBL: a large-scale bioactivity database for drug discovery.” Nucleic acids research 40.D1 (2012): D1100–D1107.

19. Sollis, Elliot, et al. “The NHGRI-EBI GWAS Catalog: knowledgebase and deposition resource.” Nucleic acids research 51.D1 (2023): D977–D985.

20. Moreno, Pablo, et al. “Expression Atlas update: gene and protein expression in multiple species.” Nucleic acids research 50.D1 (2022): D129-D140.E

21. Tian, Ruilin, et al. “Genome-wide CRISPRi/a screens in human neurons link lysosomal failure to ferroptosis.” Nature neuroscience 24.7 (2021): 1020–1034.

22. Rosonovski, Summer, et al. “Europe PMC in 2023.” Nucleic Acids Research 52.D1 (2024): D1668–D1676.

23. Food and Drug Administration Amendments Act of 2007, Pub L No. 110-85, 121 Stat 823 (2007). Accessed January 18, 2026. https://www.govinfo.gov/content/pkg/PLAW-110publ85/pdf/PLAW-110publ85.pdf

24. Buyse, Marc, et al. “Central statistical monitoring of investigator-led clinical trials in oncology.” International Journal of Clinical Oncology 25.7 (2020): 1207–1214.

25. Stoeger, Thomas, et al. “Large-scale investigation of the reasons why potentially important genes are ignored.” PLoS biology 16.9 (2018): e2006643.

26. Adda, Jérôme, Christian Decker, and Marco Ottaviani. “P-hacking in clinical trials and how incentives shape the distribution of results across phases.” Proceedings of the National Academy of Sciences 117.24 (2020): 13386–13392.

27. Montinari, Maria Rosa, Sergio Minelli, and Raffaele De Caterina. “The first 3500 years of aspirin history from its roots–A concise summary.” Vascular pharmacology 113 (2019): 1–8.

28. Tian, Shubo, et al. “Opportunities and Challenges for ChatGPT and Large Language Models in Biomedicine and Health.” arXiv preprint arXiv:2306.10070 (2023).

29. Vasan, Kishore, Deisy Morselli Gysi, and Albert-László Barabási. “The clinical trials puzzle: How network effects limit drug discovery.” Iscience 26.12 (2023).

30. Kingsmore, Stephen F., et al. “Genome-wide association studies: progress and potential for drug discovery and development.” Nature Reviews Drug Discovery 7.3 (2008): 221–230.

31. Martin, Paul, Nik Brown, and Alison Kraft. “From bedside to bench? Communities of promise, translational research and the making of blood stem cells.” Science as Culture 17.1 (2008): 29–41.

32. Wong, Chi Heem, Kien Wei Siah, and Andrew W. Lo. “Estimation of clinical trial success rates and related parameters.” Biostatistics 20.2 (2019): 273–286.

